# Patient-derived mutations impact pathogenicity of SARS-CoV-2

**DOI:** 10.1101/2020.04.14.20060160

**Authors:** Hangping Yao, Xiangyun Lu, Qiong Chen, Kaijin Xu, Yu Chen, Linfang Cheng, Fumin Liu, Zhigang Wu, Haibo Wu, Changzhong Jin, Min Zheng, Nanping Wu, Chao Jiang, Lanjuan Li

## Abstract

The sudden outbreak of the severe acute respiratory syndrome–coronavirus (SARS-CoV-2) has spread globally with more than 1,300,000 patients diagnosed and a death toll of 70,000. Current genomic survey data suggest that single nucleotide variants (SNVs) are abundant. However, no mutation has been directly linked with functional changes in viral pathogenicity. We report functional characterizations of 11 patient-derived viral isolates. We observed diverse mutations in these viral isolates, including 6 different mutations in the spike glycoprotein (S protein), and 2 of which are different SNVs that led to the same missense mutation. Importantly, these viral isolates show significant variation in cytopathic effects and viral load, up to 270-fold differences, when infecting Vero-E6 cells. Therefore, we provide direct evidence that the SARS-CoV-2 has acquired mutations capable of substantially changing its pathogenicity.

## Introduction

Severe acute respiratory syndrome coronavirus 2 (SARS-CoV-2; previously referred to as 2019-nCoV), associated with the ongoing outbreak of atypical pneumonia, has already caused a global pandemic beginning in Wuhan, Central China, despite China’s extensive systematic effort to contain the outbreak. As of April 7, 2020, SARS-CoV-2 has infected more than 1.3 million people around the world with a death toll of 70,000. The numbers are still increasing rapidly. The estimate of the incubation period of SARS-CoV-2 (mean, 5.1 days; range, 4.5 to 5.8 days) (Lauer et al., 2020) is in line with those of other known human coronaviruses, such as SARS (mean, 5 days; range, 2 to 14 days) (Varia et al., 2003) and MERS (mean, 5 to 7 days; range, 2 to 14 days) (Virlogeux et al., 2016). The reproductive number of SARS-CoV-2 is likely to be from 1.4 to 6.5, with a mean of 3.3 (Liu et al., 2020), which is slightly higher than SARS, i.e., 2-5 (Bauch et al., 2005; Lipsitch et al., 2003) and MERS, i.e., 2.7-3.9 (Lin et al., 2018). More than half of patients with SARS-CoV-2 showed no signs of fever before hospitalization (Guan et al., 2020). Strikingly, Coronavirus Diease-2019 (COVID-19) can be transmitted by asymptomatic patients, who show no fever, gastrointestinal or respiratory symptoms, and have normal chest computed tomography (Bai et al., 2020; Hu et al., 2020), making it much more challenging to prevent the spread of COVID-19. Moreover, SARS-CoV-2 can remain viable and infectious in aerosols for multiple hours and up to 7 days on surfaces (van Doremalen et al., 2020). Although multiple *in vitro* studies or clinical trials on inhibitors or drugs were carried out, no effective cures or vaccines have been found so far (Cao et al., 2020; Hoffmann et al., 2020; Wang et al., 2020). The World Health Organization (WHO) declared COVID-19 a Public Health Emergency of International Concern on 30 January 2020, and raised the threat of the SARS-CoV-2 pandemic to the “very high” level on February 28, 2020.

SARS-CoV-2 is the seventh member of enveloped RNA beta-coronavirus (Sarbecovirus subgenus) (Zhu et al., 2020); SARS-CoV-2, SARS-CoV and MERS-CoV can lead to devastating diseases, while HKU1, NL63, OC43 and 229E are related with mild symptoms (Corman et al., 2018). So far, no recombination events were detected (Yu, 2020), although this could be at least partially due to the fact that most viral isolates were sequenced with short-reads platform. The transmembrane spike (S) glycoprotein mediates viral entry into host cells through homotrimers protruding from the viral surface. The S protein includes two domains: S1 for binding to the host cell receptor and S2 for fusion of the viral and cellular membranes, respectively (Tortorici and Veesler, 2019). Both SARS-CoV-2 and SARS-CoV use the angiotensin converting enzyme 2 (ACE2) to enter target cells (Walls et al., 2020). ACE2 is expressed in human nasal epithelial cells and lung, spermatogonia, leydig, sertoli, gastric, duodenal, and rectal epithelial cells (Wang and Xu, 2020; Xiao et al., 2020; Zhao et al., 2020). The receptor binding domain (RBD) in the S protein is the most variable genomic part in the betacoronavirus group (Wu et al., 2020; Zhou et al., 2020), and some sites of S protein might be subjected to positive selection (Lv et al., 2020). Despite the abundant variability of SARS-CoV-2, one key question remains as to whether these mutations have any real functional impact on the pathogenicity of SARS-CoV-2. This is crucial in our understanding of the viral infectious mechanisms and dictates the strategy of drug and vaccine development in preparation for the next stage of the pandemic.

To address this, we characterized 11 SARS-CoV-2 viral isolates from patients admitted to Zhejiang University-affiliated hospitals in Hangzhou, China, situated 757 KMs to the east of Wuhan (see Materials and Methods). Super-deep sequencing of the 11 viral isolates on the Novaseq 6000 platform identified 1-5 mutations in the coding sequences among the viral isolates. Mixed viral populations (representing quasi-species) were also observed. We infected Vero-E6 cells with 11 viral isolates and quantitatively assessed their viral load at 1, 2, 4, 8, 24, and 48 hours post infection (P.I.) and their viral cytopathic effects (CPE) at 48 and 72 hours P.I.. Our results show that the observed mutations can have a direct impact on the viral load and CPE when infecting Vero-E6 cells, as much as 270-fold differences between the extremities. This finding suggests that the observed mutations in our study, and possibly in the viral isolates collected around the world, can significantly impact the pathogenicity of SARS-CoV-2.

## Results

### Summary of the epidemiological history of the patients

The samples of the 11 patients involved in this study were collected during the early phase of the COVID-19 breakout in China, dates ranging from 1/22/2020 to 2/4/2020 (Table 1). 10 of the 11 patients had clear connections with Wuhan city, where the SARS-CoV-2 was originally identified. 5 of the 11 people either worked in or traveled to Wuhan before they were diagnosed, and another 5 had close contact with people who lived in Wuhan, and the remaining person had contact with people who were COVID-19 victims. Notably, patients ZJU-4, −5, −9 attended the same business conference where a few colleagues from Wuhan were present. These patients therefore constitute 1st and 2nd generations of the viral victims based on their epidemiological history. The 11 patients include 8 males and 3 females, with ages ranging from 4 months to 71 years old. There are no clear criteria in selecting these patients other than the fact that they were all admitted into Zhejiang University-affiliated hospitals in Hangzhou. All except one of the patients had moderate or worse symptoms. 3 patients had co-morbidity conditions and one patient needed ICU treatment. Luckily, all of the patients have recovered as of the time of writing this article.

**Table 1.**
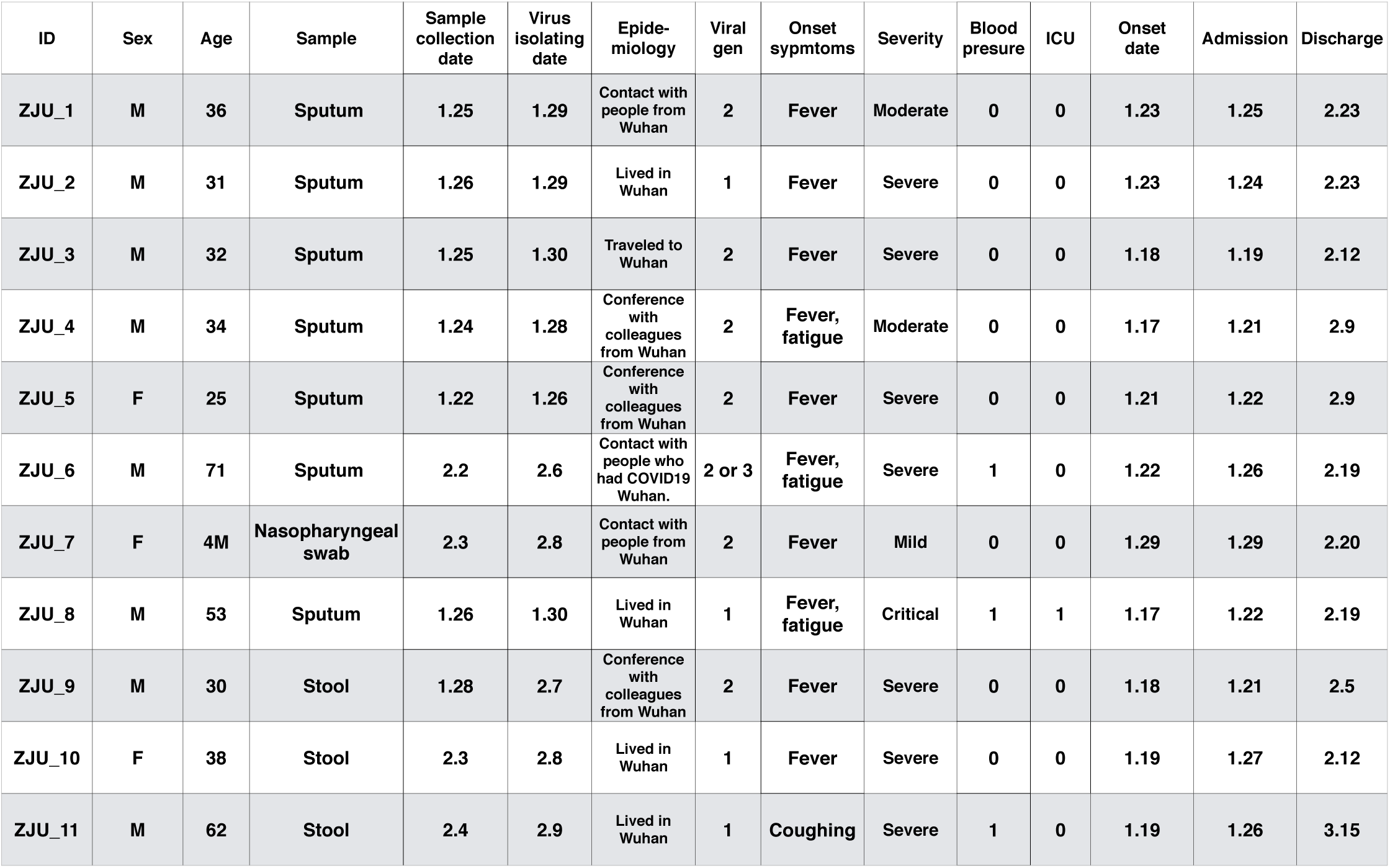
A summary of the epidemiological information of the 11 patients involved in this study. The “Viral gen” (viral generation) was inferred based on their exposure history.

### Ultra-deep sequencing reveals diverse mutations in the patient-derived viral isolates

To assess the mutational spectrum of these 11 viral isolates, ultra-deep sequencing of the isolated viral genomic RNA was performed on the Illumina Novaseq 6000 platform, generating on average 245 million post-cleaning reads/67.16 Gb per sample (Table S1; average coverage exceeding 2,000,000 X). This extraordinary depth is partially due to the small genome of the SARS-CoV-2, which enables us to identify mutations with high confidence. Moreover, in cases where the viral populations are not homogenous, the depth could help us to characterize alleles with very low frequency.

In total, 33 mutations were identified (including 10 mutations observed in mixed-populations), and 19 of these mutations were novel, according to the comparison with 1111 genomic sequences available at GISAID on 3/24/2020 (Fig. 1, S1, and S2). Specifically, G11083T and G26144T were found in ZJU-1, and both of these mutations are known as founding mutations for a large group of viruses (Capobianchi et al., 2020). C8782T and T28144C were found in two of our viral isolates, ZJU-2 and ZJU-8, and these two are known as the founding mutations for another large group of viral isolates (Capobianchi et al., 2020). Interestingly, mutation T22303G was found in five viral isolates (ZJU-2, −5, −9, −10, and −11) and ZJU-5 and ZJU-9 were exposed to the same potential source of infection during a business conference (Table 1). Previously, only one viral isolate identified in Australia had the T22303G mutation. Strikingly, the viral isolate from patient ZJU-4, who attended the same conference as ZJU-5 and ZJU-9, has a novel mutation, A22301C, which causes the same missense mutation at the protein level (S247R in the S protein) as T22303G, mutating the 1st instead of the 3rd position in the respective codon. Observations of these two single nucleotide variants can only be coincidental, albeit very unexpected. Finally, the ZJU-11 has 4 mutations in the ORF7b gene, 3 of which are consecutive and introduce 2 mutations at the protein level. Di-nucleotide and Tri-nucleotide mutations are of course rarer than SNV, but not exponentially so, according to previous mutational accumulation studies in prokaryotes (Lynch, 2007).

**Fig. 1.**
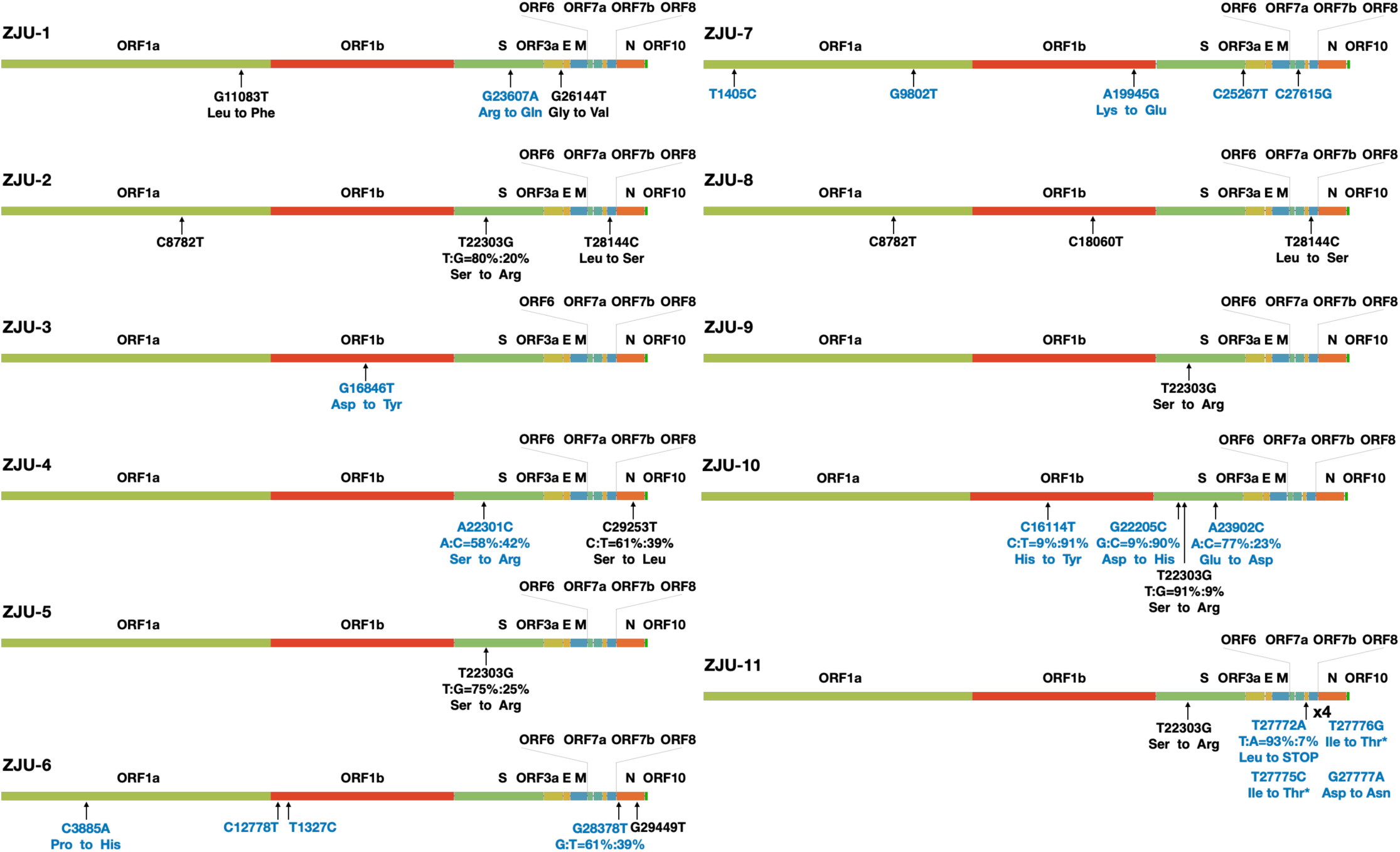
A summary of the mutations identified in each of the 11 viral isolates. Each ORF of the viral genome was denoted based on the annotations of NC_045512.2 as provided by NCBI. If a mutation was observed in the context of a mixed population, the respective percentages of the top two alleles are provided. Changes at the amino acid level are provided if applicable. Blue color indicates novel mutation at the time of writing the article.

It is important to note that while the sequence data deposited in GISAID are very helpful in tracking inter-personal variation of the virus, we still do not know much about intra-personal viral evolutionary dynamics. For example, in ZJU-4 and ZJU-10, alleles of two separate sites have very similar frequency distributions, indicating that these two sites are probably linked, representing at least two haplotypes within the viral populations. And as revealed by this study, 6 of the identified mutations would have been ignored if using the consensus sequences for analyses. Taken together, despite only 11 patient-derived isolates being analyzed in this study, we observed abundant mutational diversity, including several founding mutations for different major clusters of viruses now circulating globally. This diverse mutational spectrum is consistent with their relatively early sampling time and relative proximity to Wuhan city, where the first viral strain was identified. The full mutational diversity of the virus in Wuhan city in the early days is still unknown to this day, due to limited sampling (Lu et al., 2020; Zhou et al., 2020).

### Phylogenetic analysis of the patient-derived viral isolates reveals their diverse evolutionary history

To understand the phylogenetic context of 11 viral isolates with respect to the corpus of available SARS-CoV-2 sequencing data, we acquired 725 high quality and high coverage SARS-CoV-2 genomes from GISAID (downloaded on 3/21/2020), including the Yunnan RaTG13 viral strain and the Guangdong pangolin viral strain as the outgroup. We aligned the 736 genomic sequences with MAFFT (see Materials and Methods) and trimmed the full-length alignment with trimAL (see Materials and Methods) to remove any spurious parts of the alignment. We used iqtree (see Materials and Methods) to construct a 1000-times bootstrapped maximum-likelihood phylogenic tree of the 736 viral sequences based on 835 parsimony informative sites (Fig. 2A). The resulting phylogenetic tree is largely consistent with the phylogenetic analysis being updated on GISAID (Fig. S3). We want to emphasize that due to the rapidly developing COVID-19 situation, tens or even hundreds of new sequences are being uploaded to the GISAID every day. As a result, new observations may be generated when more data are available.

**Fig. 2.**
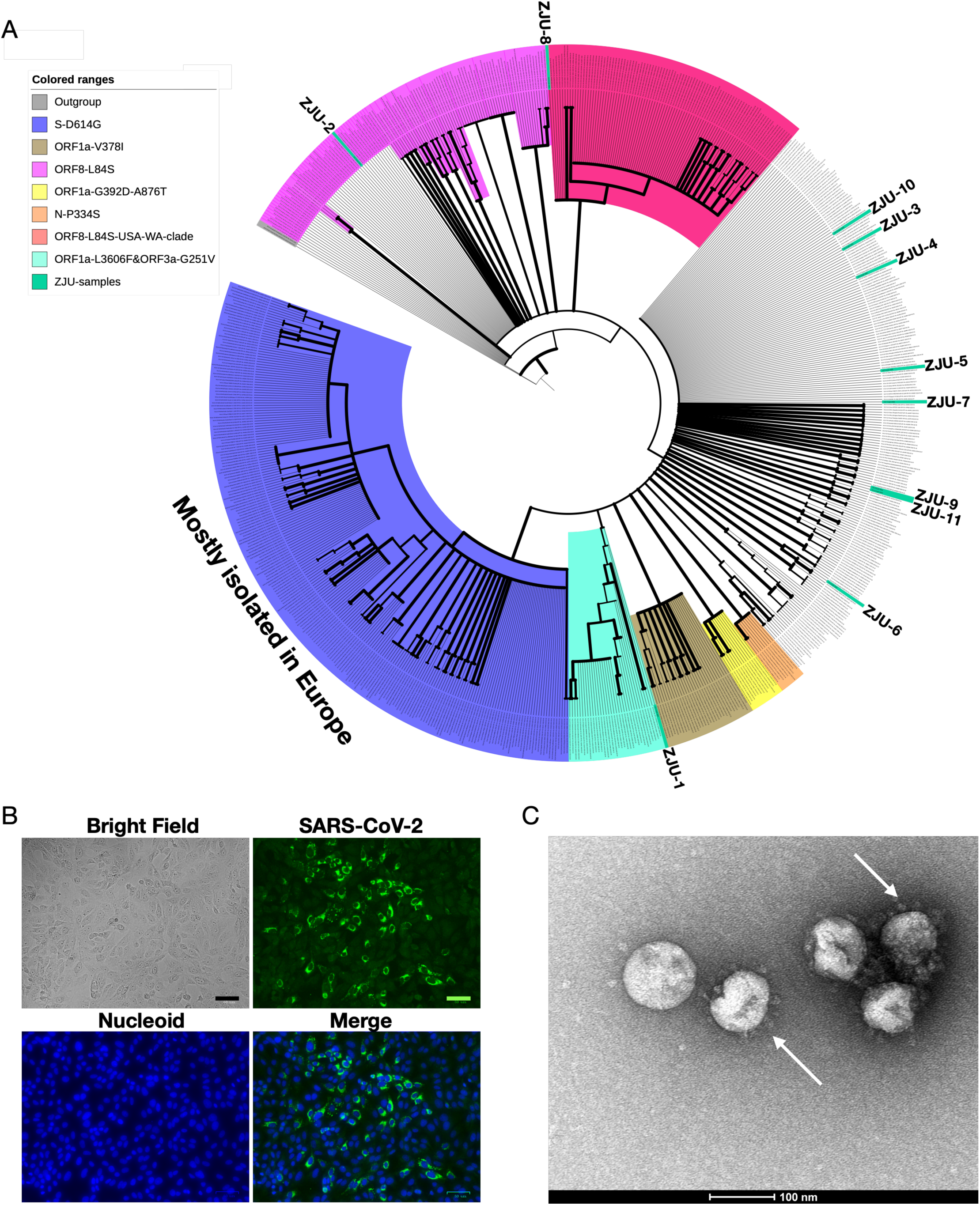
Characterizations of the patient-derived SARS-CoV-2 isolates. (A) Phylogenetic analyses of the 11 viral isolates in the context of 725 SARS-CoV-2 sequences downloaded from GISAID. The 1000-times bootstrapped maximum likelihood tree was constructed to demonstrate the phylogenetic context of the 11 viral isolates. Major and minor clusters were color-coded and denoted as shown in the “colored ranges” inset box. All ZJU-samples were color-codded as green. The width of a branch indicates bootstrap supporting level. (B) Fluorescent labeling of the viral S protein indicates that isolated SAR-CoV-2 viral particles (Green) bind to the peripherals of the Vero-E6 cells (DNA stained as Blue) prior to entry. Scale bars, 50 µm. (C) A representative TEM picture of the isolated SAR-CoV-2 viral particles, arrows indicate the iconic “crown” consisted of S proteins (Spike). Scale bar, 100 nm.

We observed quite a few sets of founding mutations. Specifically, we note the following three biggest clusters in our phylogenetic analysis: 1. Three nucleotide mutations C241T (silent), C14408T (silent), and A23403G (D614G in S) found a group of 231 viral sequences (Fig. 2A; S-D614G cluster), most of which were isolated in Europe; 2. Two nucleotide mutations C8782T (silent) and T28144C (L84S in ORF8), found a group of 208 viral sequences (Fig. 2A; ORF8-L84S cluster), which is not monophyletic in our analysis (Fig. 2A and S3). However, a distinct monophyletic subclade of 92 sequences within the ORF8-L84S cluster can be observed, mainly composed of viral sequences isolated from Seattle, USA (Fig. 2A; ORF8-L84S-USA-WA-clade); 3. Two nucleotide mutations, G11083T (L3606F in ORF1a), and G26144T (G251V in ORF3a) found a group of 34 viral sequences, most of which were from Netherlands and England. Several smaller monophyletic clusters, defined by different sets of founding mutations (bootstrap supporting value >95), can be observed. For examples: 1. the G1937A (V378I in ORF1a) mutation founds a cluster of 31 viral sequences; 2. the G1440A and G2891A mutations, resulting in G392D and A876T mutations in the ORF1a gene, founds a cluster of 12 viral sequences, mostly from Germany or Netherlands; 3. the C15325T and C29303T mutations, resulting in P344S mutation in the N gene, founds a small cluster of 8 sequences, all of which are from China or Japan. When integrating the characterized 11 viral isolates into the phylogenetic analysis, they are dispersed across the entire phylogenetic space. ZJU-1 clusters with the ORF1a-L3606F & ORF3a-G251V groups, as it has both of the two defining mutations (Fig. 2A). ZJU-2 and ZJU-8, on the other hand, cluster with the ORF8-L84S cluster because they both have the two founding mutations (Fig. 2A). ZJU-9 and ZJU-11 cluster with an Australian isolate because of the aforementioned T22303G mutation. The rest of the group either have few mutations or novel mutations that do not cluster with any known sizable groups, reflecting the extensive diversity within our 11 samples. Taken together, some monophyletic clusters of viruses do show obvious geographic patterns (Europe and WA-USA especially), but this could be due to the founding effect of respective mutations that happened early during the initial phase of the pandemic.

### Patient-derived SARS-CoV-2 isolates show significant variation in viral copy number and cytopathic effects when infecting Vero-E6 cells

There is much speculations and many theories behind the observed mutations in sequenced viral isolates. Theoretically, one usually needs not often invoke selection arguments in explaining the origin of these mutations, as the human to human infection process is a series of repeated naturally-occurring bottlenecking events, in which the seeding viral population can be as little as hundreds of viral copies (Forni et al., 2017). Therefore, a significant portion of the genetic diversity or even population-specific fixations could be due to this process, where selection plays a small role (Renzette et al., 2017). We conducted Tajima’s test of neutrality using the constructed alignment of viral sequences and Taijima’s D is −2.8874 with a nucleotide diversity (π) of 0.000641 (p < 0.05 according to simulations performed in (Tajima, 1989), indicating that the SARS-CoV-2 genome has an excess of low-frequency alleles due to recent population expansions, consistent with the repeated bottlenecking events during viral infections. However, certain mutations do provide selection advantages or disadvantages under specific circumstances, as shown by the discovery that adaptive mutations are highly enriched in the interface between the S protein and the human ACE2 receptor (Ou et al., 2020).

To examine the mutational impact of the patient-derived SARS-CoV-2 isolates, we conducted *in vitro* infectivity assay. We chose *in vitro* assay because COVID-19 patients show a wide variety of clinical symptoms ranging from asymptotic to death, and epidemiological research have shown that the clinical outcomes are heavily influenced by individual’s age, complications, and other potential unknown parameters (Guan et al., 2020). We first examined whether the viral isolates could successfully bind to Vero-E6 cells as expected (Fig. 2B), and visually identified the viral particles with the iconic “crown” formed by S proteins (Fig. 2C and S4A). We then infect the Vero-E6 cells with all 11 patient-derived viral isolates and harvest the cells at 1, 2, 4, 8 (in quadruplicates), 24, and 48 (in duplicates) hours P.I. (see Materials and Methods); we included the supernatant because cell death releases viral particles. DIC micrographs of the cells at 48 hours and 72 hours P.I. were also taken to assess the CPE. We used specific real-time reverse transcriptase–polymerase chain reaction (RT-PCR) targeting ORF1a, E, and N genes to detect the presence of SARS-CoV-2 (see Materials and Methods). Cycle threshold values, C_t_, were used to quantify the viral load, with lower values indicating higher viral load. Because the results based on the three genes are highly consistent (R > 0.99, p < 2.2e-16), we will only discuss the results of the ORF1a gene (Fig. 3A). We failed to detect any significant signals from our negative controls, hence we simply assigned a C_t_ value of 40 for all them.

**Fig. 3.**
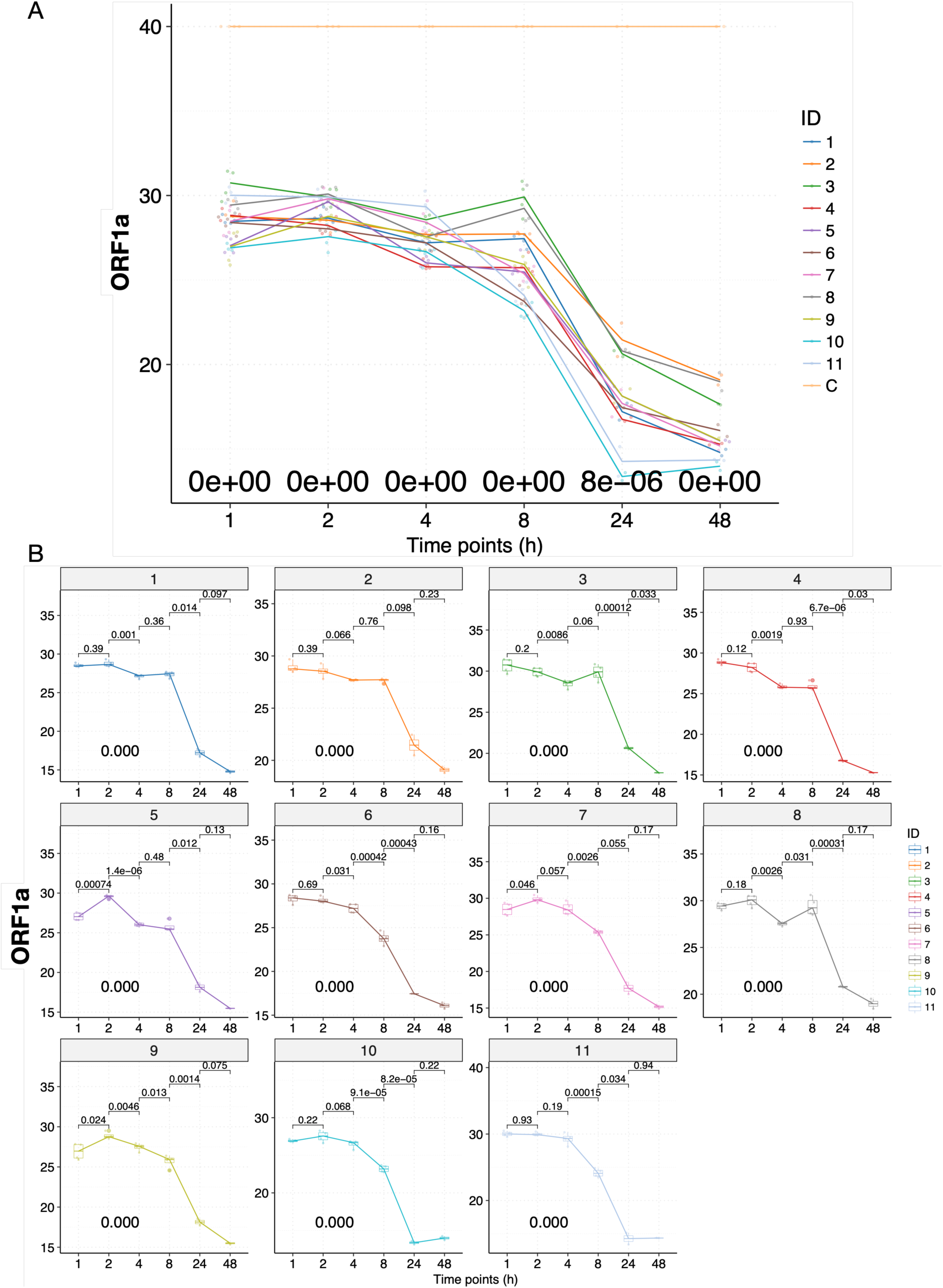
The infectivity assay reveals temporal variation in the viral load of the patient-derived SARS-CoV-2 isolates when infecting Vero-E6 cells. (A) Time-series plots of the C_t_ values (corresponding to the multiplicative inverse of viral load) of the SAR-CoV-2 ORF1a gene over the course of infectivity assay. Each viral isolate plus the negative control “C” was color-coded accordingly. P-values were calculated using the ANOVA method to compare the means of all 11 viral isolates at each time point, excluding the negative control “C”. (B) Time-series plots of the C_t_ values of the SAR-CoV-2 ORF1a gene for each of the 11 patient-derived viral isolates. P-values were calculated between consecutive time points using the t-test and adjusted p-values are shown.

Briefly, the C_t_ values of samples remained mostly flat with small fluctuations for all of the viral isolates at 1, 2, and 4 hours P.I. (Fig. 3A and B). During these early hours, viral particles are binding to gain access into the cells, and replications would rarely occur (Schneider et al., 2012). At 8 hours P.I., we observed significant decreases in Ct value (increases in viral load) for ZJU-6, ZJU-7, ZJU-9, ZJU-10, and ZJU-11. At 24 hours P.I., we observed significant decreases in C_t_ values for all of the viral isolates except for ZJU-2 and ZJU-7, although some of the viral isolates, namely ZJU-10 and ZJU-11, decreased much faster than the others (Fig. 3A and B). At 48 hours P.I., we observed small decreases for all viral isolates except for ZJU-10 and ZJU-11, both of which had presumably already plateaued at 24 hours P.I. (Fig. 3A and B). Notably, at 24 hours P.I., ZJU-2 and ZJU-8, members of the ORF-8-L84S cluster (majority of USA WA-Seattle isolates are in this group), showed considerably lower viral loads (Fig. 4A). On the other hand, ZJU-1, which clusters with the S-D614G clade (mostly found in Europe), has a viral load 19 times (2^4.25^) higher than ZJU-2 and ZJU-8 (Fig. 4A). In addition, a near 270-fold difference (2^8.09^) in viral load was observed between ZJU-10 and ZJU-2 at 24 hours P.I. (Fig. 4A). These differences became statistically significant at 48 hours P.I., and are reproducible when analyzing data on gene E and N (Fig. S4B and C; Fig. S5A). Therefore, different viral isolates, which are defined by different mutations in their genomes, exhibit a significant variation of viral load when infecting Vero-E6 cells.

**Fig. 4.**
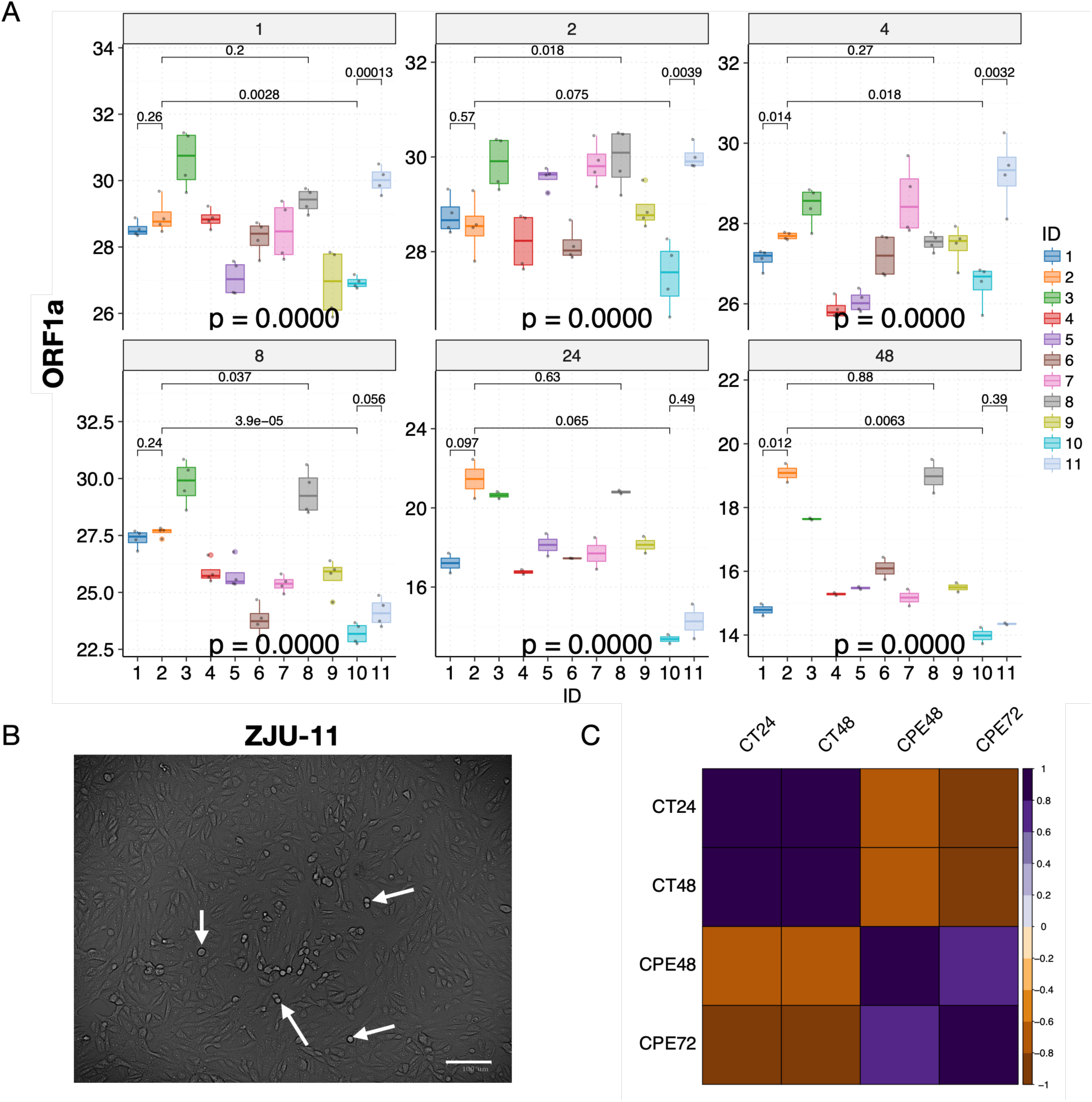
The changes in CPE and viral load are highly correlated. (A) Significant variations in viral load were observed at each time point. Mean C_t_ values of selected viral isolates are displayed and color-coded, respectively. P-values were calculated using the ANOVA method to compare the means of all 11 viral isolates at each time point. Pair-wise p-values were calculated between isolates using the t-test and adjusted p-values are shown. (B) Cytopathic effects were visible under the microscope 48 hours P.I., white arrows indicate representative cells undergoing lysis. (C) Cytopathic effects (CPE) were highly correlated with viral load (CT) in viral infectivity assay. Pearson correlations were calculated and p-values were adjusted accordingly; only correlations with adjusted p-value < 0.05 are shown. Note that the CT values are negatively correlated with CPE values because CT values represent the inverse number of viral loads.

We next examined whether a higher viral load leads to more cell death (Fig. 4B). When examining these cell lines under a microscope at 48 hours and 72 hours P.I., the CPE, or the cell death rate, are highly consistent with the viral load data (Fig. 4C and S5B, C, and D; at 48 hours P.I., C_t_ vs CPE, R = −0.72, p = 0.015), indicating that a higher viral load leads to a higher cell death ratio. Note that the C_t_ numbers are negatively correlated with the CPE because a lower C_t_ number means a higher viral load.

## Discussion

The quickly-developing COVID-19 pandemic has already infected millions of victims and caused 70,000 deaths globally. While many ongoing research projects are attempting to track the evolutionary origin of the virus, find the mechanisms of infection, and produce vaccines or drugs against the virus, we sought to establish the genotype-phenotype link behind the abundant diversity being observed as a result of global sequencing efforts (GISAID). Due to the extremely wide variety of clinical symptoms shown in the patients, establishing a genotype-phenotype link in patients would be very difficult. The *in vitro* cell line provides an ideal system to examine the mutational impact of different isolates of viruses, when all other confounding factors are removed. Although the Vero-E6 cell line was not derived from human, the ACE2 protein of the Vero-E6 cell line is highly similar to that of Human (Fig. S6) and we provided direct evidence that the SARS-CoV-2 can infect the cell line (Fig. 2B).

Several findings stand out in our study: 1. A diverse collection of mutations was identified in the 11 viral isolates, including two sets of founding mutations for two major clusters of viruses currently infecting the world population. In addition, 19 of the 31 identified mutations are novel, despite the relatively early sampling dates, indicating that the true diversity of the viral strains is still largely underappreciated; 2. remarkably, the T22303G and A22301C mutations result in the same S247R mutation in the S protein (Fig. 1 and S1), mapping to the existing structure revealed that this residue is located in a flexible loop region within the N-terminal domain of the S1 subunit of S protein, although the exact position of S247 could not be determined (Fig. S7, red arch). While the N-terminal domain is not directly involved with binding to ACE2 (Walls et al., 2020) we note that this domain is positioned right next to the C-terminal domain, which binds to ACE2. Interestingly, the T22303G mutation was observed in 5 viral isolates, albeit in different proportions, indicating that this specific mutation was already present in the early days of pandemic, and probably in a significant number of people of Wuhan, despite the fact that it is still largely missing from current GISAID collection. This could be due to the founding effect of mutations, in which case the T22303G mutation was not transmitted out of the China during the early days; 3. The tri-nucleotide mutation in ZJU-11 is unexpected; we note that this specific viral isolate is quite potent in our viral load and CPE assay, and its patient remained positive for an astounding period of 45 days and was only recently discharged from the hospital (Table 1). Investigating the functional impact of this tri-nucleotide mutation would be highly interesting. We note that in the current database, another trinucleotide mutation (G28881A, G2882A and G28883C) has been identified, which also results in two missense mutations at the protein level (Fig. S8). It leads to a cluster of more than 300 viral strains as of the time of writing this article, and its mutational impact on the viral pathogenicity would be worth investigating. Finally, in contrary to the recent report that a viable viral isolate could not be obtained from stool samples, three of our viral isolated were extracted from stool samples, indicating that the SARS-CoV-2 is capable of replicating in stool samples (Woelfel et al., 2020).

In short, our study provides direct evidence that mutations currently occurring in the SARS-CoV-2 genome have the functional potential to impact the viral pathogenicity. Therefore, viral surveillance should be also performed at the cellular level when possible, in addition to the accumulating genomic sequencing data. Furthermore, characterizations of all founding mutations in the major geo-based clusters of viruses could be very useful in helping determining if there are actionable pathogenicity differences to aid the current battle against the virus. Finally, similar to flu, drug and vaccine development, while urgent, need to take the impact of these accumulating mutations, especially the founding mutations, into account to avoid potential pitfalls.

## Data Availability

The full-genome sequences of the 11 viral isolates have been deposited to the GISAID collection with the following IDs: EPI_ISL_415709, EPI_ISL_416042, EPI_ISL_416044, EPI_ISL_416046, EPI_ISL_415711, EPI_ISL_416047, EPI_ISL_416425, EPI_ISL_416473, EPI_ISL_416474, EPI_ISL_418990, and EPI_ISL_418991.

## STAR Methods

### Experimental Model and Subject Details

Patients with confirmed COVID-19 were admitted in the First Affiliated Hospital from Jan 19 to Mar 5, 2020. The First Affiliated Hospital, located in Hangzhou, Zhejiang Province, China, is one of the major provincial hospitals designated to receive patients with COVID-19 infection across the Zhejiang Province; therefore, patients with severe symptoms outside of Hangzhou were also admitted. Starting Jan 10, 2020, all patients presenting to the hospital’s fever clinic were screened by clinical staff for COVID-19 infection utilizing criteria for suspected cases as defined by the National Health Commission of China’s clinical diagnosis and management guideline for COVID-19 (China National Health Committee, 2020). Briefly, patients were screened based on their clinical symptoms and their risk of epidemiological exposure, including past travel to Hubei Province or close contact with people who had visited Hubei Province during the COVID-19 outbreak. As the pandemic continued to spread, the probability of transmission outside of Hubei Province increased. The epidemiological exposure to Hubei Province was not a prerequisite for suspected cases. All suspected cases were determined by laboratory tests and based on positive results of qRT-PCR assay for COVID-19. Patients were excluded if two qRT-PCR tests 24 hours apart both suggested negative results. Patients’ clinical samples which PCR test C_t_ value less than 28 were collected to isolate SARS-Cov-2.

## Method Details

### Sample collection, Viral isolation, cell infection, and electron microscopy

All samples, sources including sputum, nasopharyngeal swab, and stool, were collected from patients with COVID-19 with consent from all patients. The study was approved by the Clinical Research Ethics Committee of The First Affiliated Hospital, School of Medicine, Zhejiang University (Approval notice 2020-29) for emerging infectious diseases. All collected samples were sent to BSL-3 lab for viral isolation within 4 hours.

The sputum, stool, and nasopharyngeal swab samples were pre-processed by first mixing with appropriate volume (Sputum, 5-10 volumes; Stool, 2 ml/100 mg; Nasopharyngeal swab, 1 volume) of MEM medium with 2% FBS, Amphotericin B (100 ng/ml), Penicillin G (200 units/ml), Streptomycin (200 µg/ml), and TPCK-trypsin (4 µg/ml). The supernatant was collected after centrifugation at 3000 rpm at room temperature. Before infecting Vero-E6 cells, all collected supernatant was filtered using a 0.45 µm filter to remove cell debris etc.

For viral infection and isolation, 3 ml of filtered supernatant was added to Vero-E6 cells in a T25 culture flask. After incubation at 35°C for 2h to allow binding, the inoculum was removed and replaced with fresh culture medium. The cells were incubated at 35°C and observed daily to evaluate cytopathic effects (CPE). The supernatant was tested for SARS-CoV-2 by qRT-PCR (see below for qRT-PCR protocol). Once the qRT-PCR test shows positive (typically after 4-5 days of incubation), the viral particles were collected from culture supernatant by ultra-speed centrifugation (100,000x g for 2 hours) for downstream sequencing, infectivity assay, and were observed under 200 kV Tecnai G2 electron microscope.

### Immunofluorescence staining

Vero-E6 Cells were infected by SARS-CoV-2 for 24 hours, and then fixed in 80% acetone (chilled at −20°C) at room temperature for 10 min. The cells were washed three times with ice-cold PBS, blocked with 1% BSA for 30 min, and incubated with SARS-CoV-2 Spike rabbit monoclonal antibody (dilution ratio 1:200) at room temperature for 1 hour. The cells were again washed three times in ice-cold PBS, and then stained with the Alexa Fluor488®-conjugated Goat Anti-rabbit IgG secondary antibody (Abcam, Cat No. ab150077) for 1 hour at room temperature in the dark. The cells were washed three times and then incubated in 0.5 µg/mL DAPI (nuclear DNA stain) for 5 min. Immunofluorescence was detected and picture were taken using the IX81 Olympus microscope equipped with a fluorescence apparatus.

### Viral infectivity assay

Vero-E6 cells were grown in a 24-well plate and infected with different SARS-CoV-2 isolates in duplicates at MOI of 0.5. The inoculum was removed at 1 hours P.I. for the 1-hour timepoint group and at 2 hours P.I. for other timepoint groups. After incubation, the cultures were rinsed with PBS for three times and replenished with 1mL fresh culture medium. Then, the cultures were subjected to freezing immediately at −80°C for the 1- and 2-hours samples, or continued to grow for the other groups (4, 8, 24 and 48 hours) until harvest. Finally, all frozen samples from each timepoint were thawed together and the viral nucleic acid abundance was measured with SARS-CoV-2 qRT-PCR Kits, targeting ORF1a, E, and N genes (Liferiver Biotech, Shanghai). Results from the first two time points reflect the capacity of viral attachment or entry into the target cells, while results from the latter four time points represent the viral replication dynamics.

### Cytopathic effect (CPE) evaluation

Vero-E6 cell monolayers were grown and infected by different patient-derived SARS-CoV-2 isolates as described in the viral infectivity assay. At 24, 48, and 72 hours P.I., virus induced cytopathic effects were observed with a digital microscope (Bio-Rad) and pictures were taken. No obvious CPE was observed at 24 hours P.I.. Pictures taken at 48 and 72 hours P.I. were evaluated first by expert opinions and then quantitated by cell death ratio.

### Sequencing library construction

The total RNA in each deactivated viral sample was extracted using a viral RNA mini kit (Qiagen, Germany). The sequencing library was constructed using the Total RNA-Seq Kit (Kapa, Switzerland) and deep-sequenced on the illumina Novaseq 6000 platform (2 × 151 bases; Illumina Inc., San Diego, CA) by BGI genomics.

## Quantification and Statistical Analysis

### Statistical Analyses and visualization

The majority of statistical analyses and visualizations were done in Rstudio and R (at the time of writing, 1.0143 for Rstudio and 3.4.0 for R), with necessary aid from customized python scripts (2.7.4) and shell scripts (Linux). The primary R packages are mostly maintained by the Bioconductor project (https://www.bioconductor.org/, along with all their dependencies). The essential ones used are ggplot2 (2.2.1), reshape2 (1.4.3), RColorBrewer (1.1-2), scales (0.5.0), corrplot (0.84), Hmisc (4.1-1), ggrepel (0.7.0), cluster (2.0.6), factoextra (1.0.5), plyr (1.8.4), dplyr (0.7.4), psych (1.7.8), devtools (1.13.4), ggpubr (0.1.6), tidyverse (1.2.1), gridExtra (2.3), ggsci (2.8), ggbeeswarm (0.6.0), ggpmisc (0.2.16), colorspace (1.3-2).

In general, parametric statistical tests (t-test, Anova, and Pearson correlation) were used when the data distribution conforms to normality distribution (such as qPCR measurements), and non-parametric statistical tests (Wilcoxon test, Kruskal-Wallis, and Spearman correlation) were used when datasets do not conform to the normality assumption. We adjust the p values using the Benjamini & Hochberg (BH) method (Benjamini and Yekutieli, 2001) to control for False Discovery Rate (FDR), when multiple comparisons are concerned, including p value matrix constructed when calculating correlations matrix among different features or samples.

The 3D structure of the S protein was visualized and downloaded from https://www.rcsb.org/3d-view/6VSB/1.

### Sequence data processing, de novo assembling, and mutation identifications

Sequencing data was generated from Novaseq 6000 and first filtered of low quality and high barcode contamination by Soapnuke and then mapped to 43 complete genome references of 2019-nCoV (SARS-CoV-2) by BWA-MEM (Li and Durbin, 2009). References of SARS-CoV-2 were downloading from NCBI on date February 28th, 2020.

Further, mapping reads that longer than 100nt were extracted for de novo assemby by SPAdes (Bankevich et al., 2012) (v3.1.3) using an iterative short-read genome assembly module for pair-end reads. K-values were selected automatically at 33nt, 55nt and 77nt for these samples. After assembling, contigs was blasted to nt database (20190301) to confirm their origins, and only contigs belonging to coronavirus were retained for base correction. Next, filtering reads of each sample were mapped back to retained assembled contigs and bam-readcount was applied (--min-mapping-quality=5, other parameter was set default) to calculate the base frequency of every post of each assemble contigs. Meanwhile, Haplotypecaller of gatk was applied to call snp/indel based on the assembled contigs with reads quality higher than 20. Finally, bam files were inspected in igv manually to verify each mutation based on the number of reads mapped, the balance between reads mapped to plus and minus strands of the reference genome, and the relative positions of the mutations on these reads.

### Phylogenetic analysis

We acquired 725 high quality and high coverage SARS-CoV-2 genomes from GISAID (downloaded on 3/21/2020), including the Yunnan RaTG13 viral strain and the Guangdong pangolin viral strain as the outgroup. We aligned the 736 genomic sequences with MAFFT (Katoh and Standley, 2013) with options --thread 16 –globalpair --maxiterate 1000 and trimmed the full-length alignment with trimAL (Capella-Gutiérrez et al., 2009) using the -automated1 option to remove any spurious parts of the alignment, which could introduce noise to the phylogenetic analysis process. We used iqtree (Nguyen et al., 2015) with options -bb 1000 -alrt 1000 -nt 64 -asr to construct a 1000-times bootstrapped maximum-likelihood phylogenic tree of the 736 viral sequences based on 835 parsimony informative sites. The resulting phylogenetic tree was imported in iTOL and visualized (Letunic and Bork, 2011). We conducted Tajima’s test of neutrality based on the constructed alignment of viral sequences using MEGA 7 (Kumar et al., 2016).

## Supplementary Figure Legends

**Table S1.** A summary of the sequencing statistics of the 11 viral isolates involved in the study. Note that ZJU_10 and ZJU_11 were sequenced in a different batch.

| ID | Raw reads | Clean reads | Raw bases(G) | Clean bases(G) | coverage | Clean rate | Error_rate_fq<br>1 | Error_rate_fq<br>2 |
| --- | --- | --- | --- | --- | --- | --- | --- | --- |
| ZJU_1 | 239,755,367 | 227,931,734 | 71.93 | 63.96 | 2,138,916 | 88.92% | 0.04% | 0.04% |
| ZJU_2 | 211,974,282 | 195,211,677 | 63.59 | 53.28 | 1,781,761 | 83.78% | 0.04% | 0.04% |
| ZJU_3 | 421,726,717 | 378,718,257 | 126.52 | 103.91 | 3,474,902 | 82.13% | 0.04% | 0.04% |
| ZJU_4 | 485,306,221 | 434,439,201 | 145.59 | 118.99 | 3,979,199 | 81.73% | 0.04% | 0.04% |
| ZJU_5 | 232,311,525 | 205,222,721 | 69.69 | 56.11 | 1,876,400 | 80.52% | 0.04% | 0.04% |
| ZJU_6 | 342,183,998 | 273,708,578 | 102.66 | 70.92 | 2,371,668 | 69.09% | 0.04% | 0.04% |
| ZJU_7 | 227,769,540 | 191,916,976 | 68.33 | 52.18 | 1,744,975 | 76.37% | 0.04% | 0.05% |
| ZJU_8 | 355,648,629 | 331,651,060 | 106.69 | 90.7 | 3,033,140 | 85.01% | 0.04% | 0.04% |
| ZJU_9 | 287,524,792 | 260,634,803 | 86.26 | 72.7 | 2,431,194 | 84.28% | 0.04% | 0.04% |
| ZJU_10 | 136,595,606 | 101,832,137 | 40.98 | 28.24 | 944,387 | 68.91% | 0.02% | 0.03% |
| ZJU_11 | 121,791,338 | 96,729,474 | 36.54 | 27.76 | 928,335 | 75.98% | 0.02% | 0.03% |
| Average | 278,417,092 | 245,272,420 | 83.53 | 67.16 | 2,245,898 | 79.70% | 0.04% | 0.04% |

**Fig. S1.**
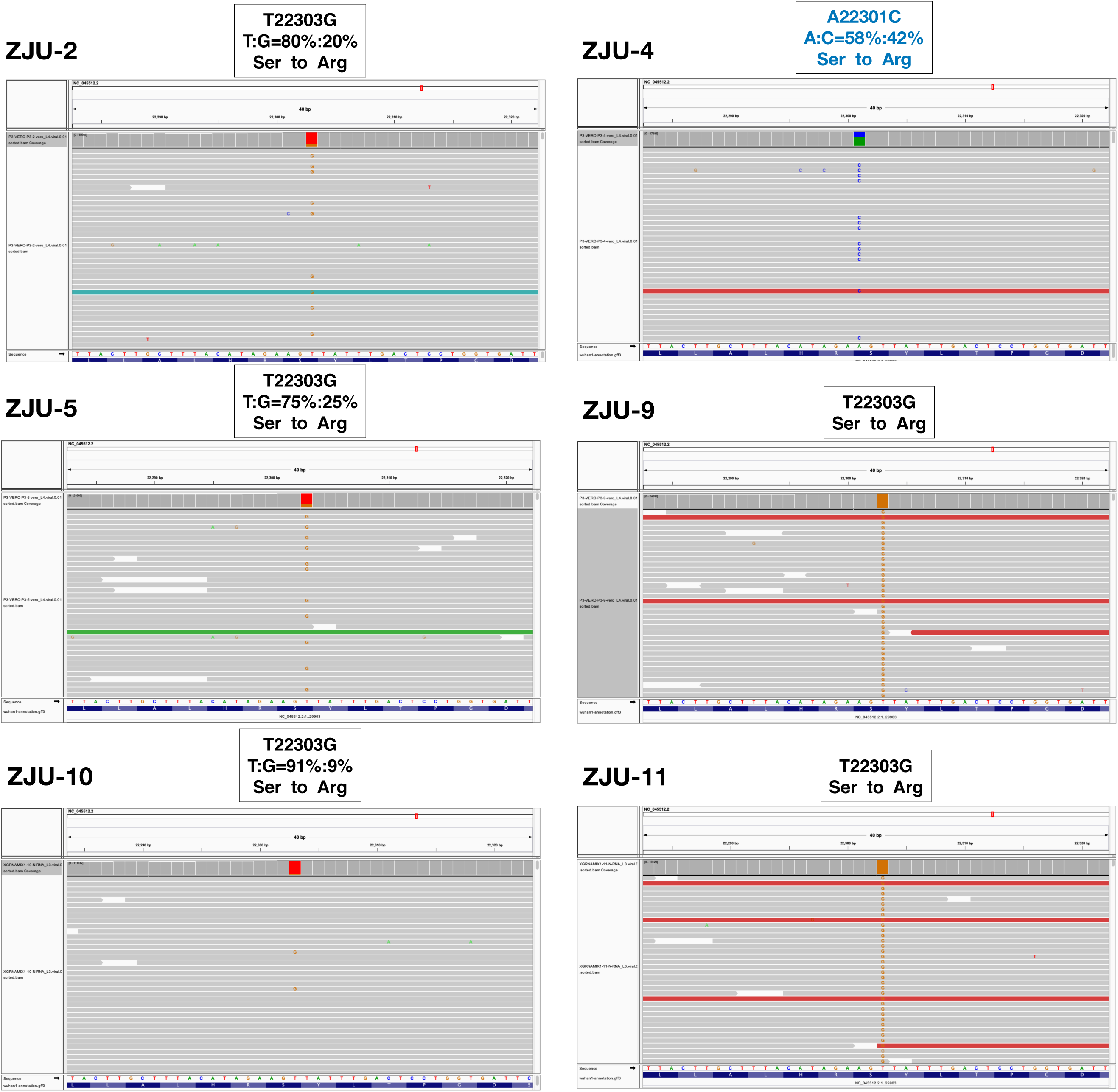
A summary of the nucleotide mutations that lead to the S247R mutations observed in the 11 patient-derived isolates. Note that some of the mutations are in the form of minor alleles. Images were produced by IGV.

**Fig. S2.**
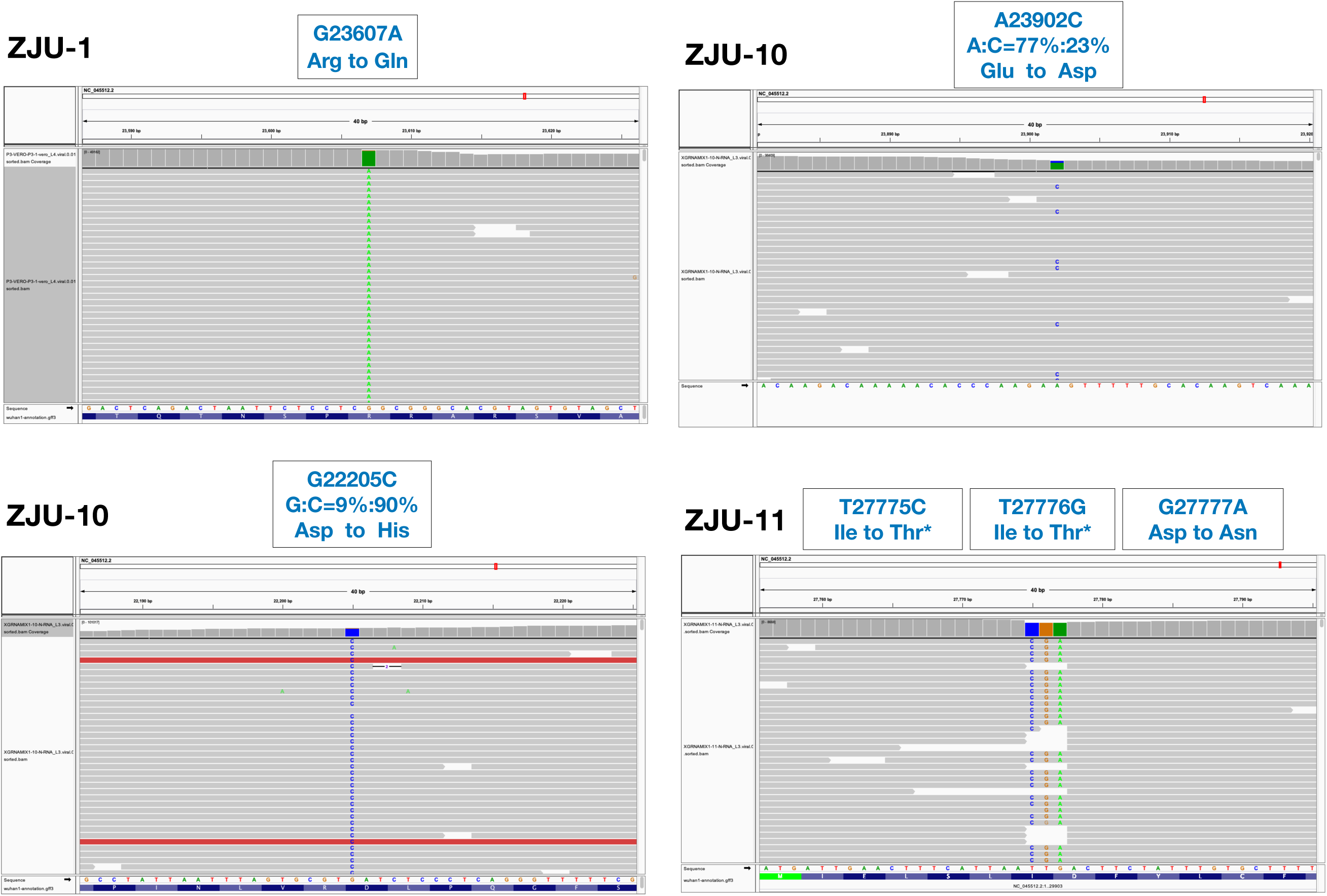
A summary of additional mutations in the S gene and the tri-nucleotide mutation. Note that some of the mutations are in the form of minor alleles. Images were produced by IGV.

**Fig. S3.**
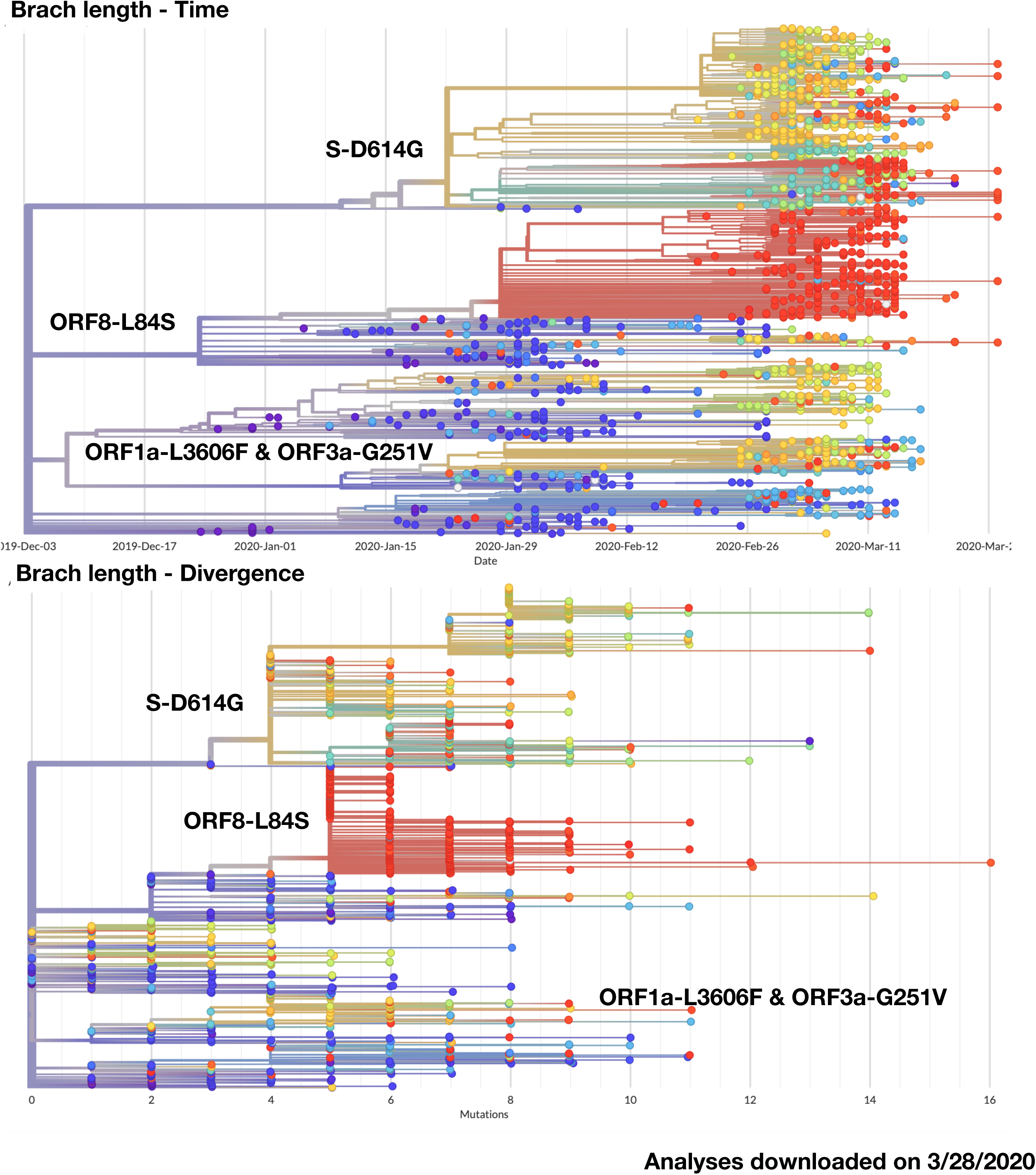
Phylogenetic analyses produced from GISAID using time (top) or number of mutations (bottom) as the branch length. Note that all three major clusters described in the study are labeled accordingly. The major distinction is that the ORF8-L84S clade is not monophyletic in our more computationally intensive and bootstrapping-supported approach.

**Fig. S4.**
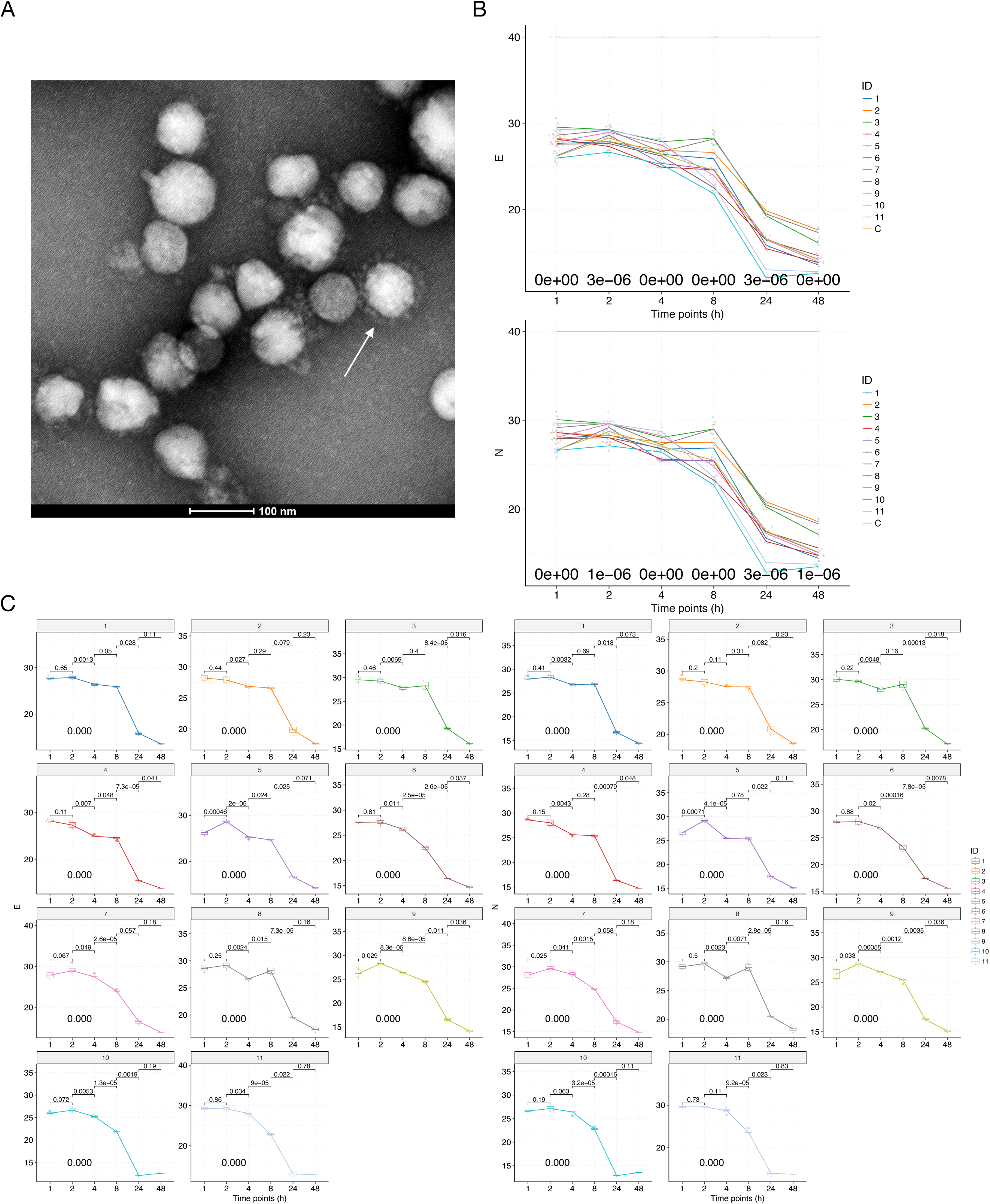
The characterizations of the 11 viral isolates. (A) A representative TEM picture of the isolated SAR-CoV-2 viral particles, arrows indicate the iconic “crown” consisted of S proteins (Spike). (B) Time-series plots of the C_t_ values (corresponding to the multiplicative inverse of viral load) of the SAR-CoV-2 E gene (top) and N gene (bottom) over the course of infectivity assay. Each viral isolate plus the negative control “C” was color-coded accordingly. P-values were calculated using the ANOVA method to compare the means of all 11 viral isolates at each time point, excluding the negative control “C”. (C) Time-series plots of the C_t_ values of the SAR-CoV-2 E gene (left) and N gene (right) for each of the 11 patient-derived viral isolates. P-values were calculated between consecutive time points using the t-test and adjusted p-values are shown.

**Fig. S5.**
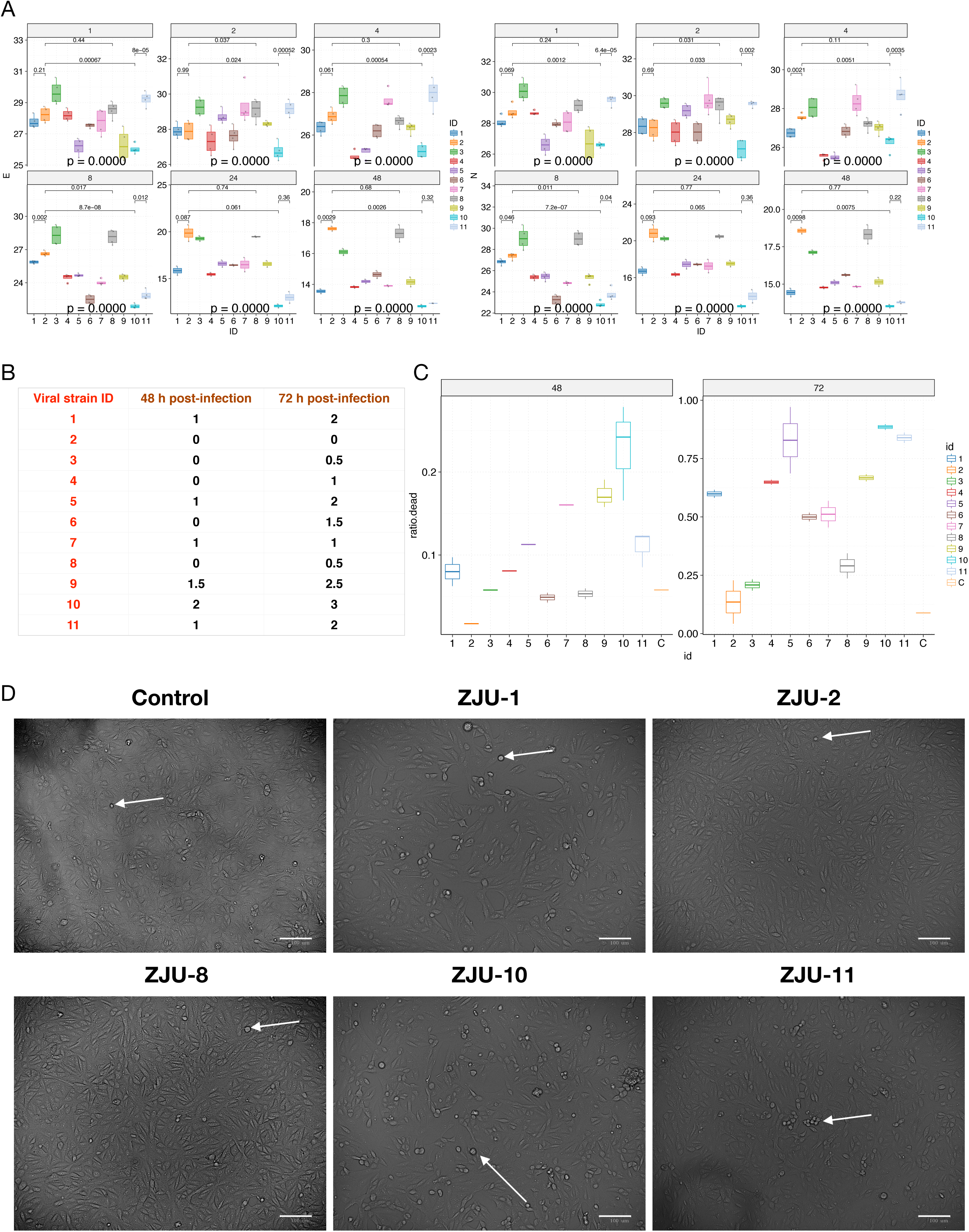
Significant variations were observed in viral load and viral CPE among the 11 patient-derived isolates. (A) Significant variations in viral load can be observed based on E gene (left) and N gene (right). (B) CPE at 48h and 72h P.I. as evaluated by an expert’s opinions. (C) CPE at 48h and 72h P.I. evaluated by quantitively calculating the cell death ratio (ratio.dead) for 1-3 images per viral isolate. The results from (B) and (C) are highly correlated (R > 0.89, p < 0.001). (D) Representative images used for CPE evaluation, arrows indicate cells facing immediate death. Scale bars, 100 µm.

**Fig. S6.**
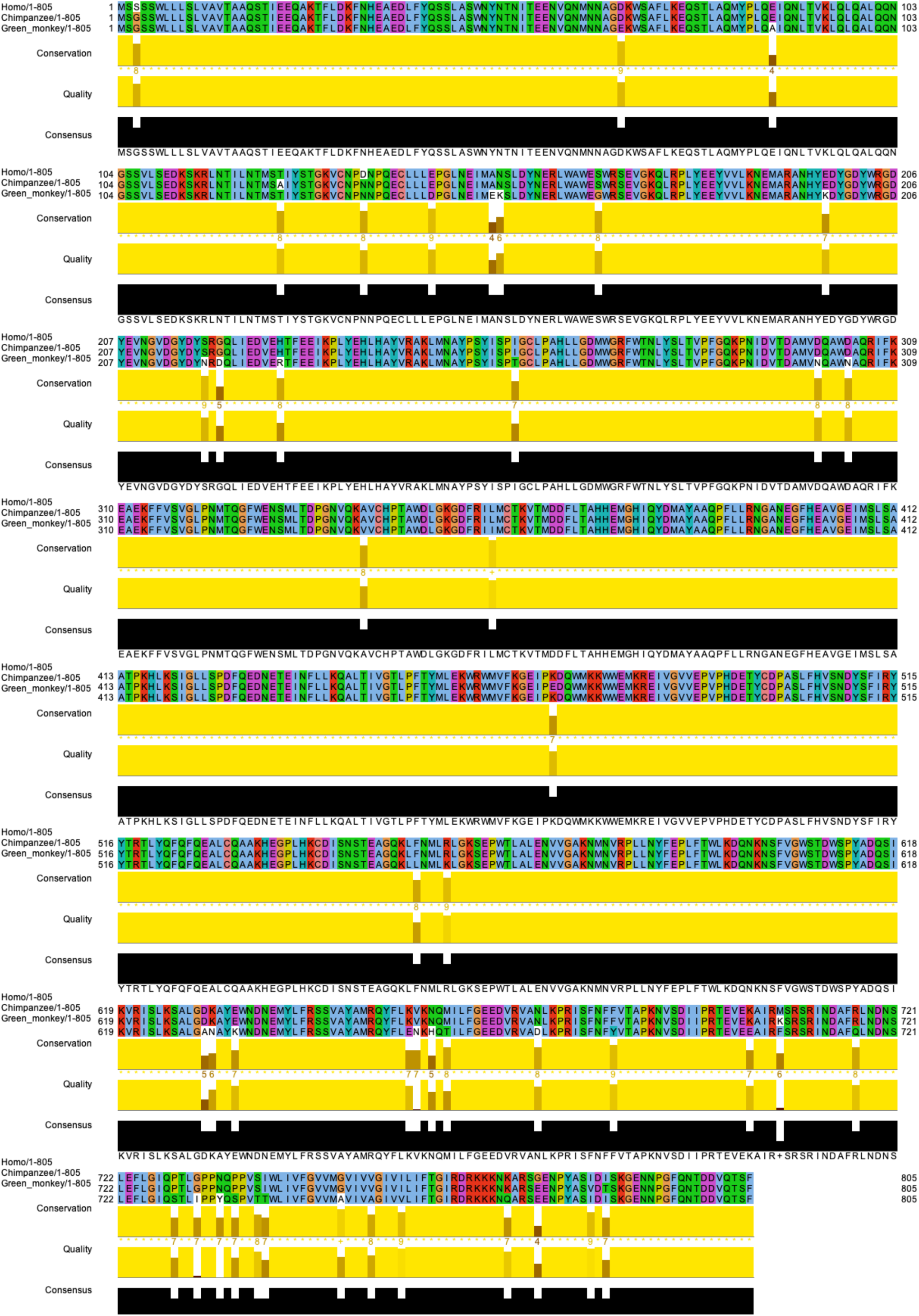
The alignment of ACE2 protein sequences from human (Homo), Chimpanzee, and green monkey (from which the Vero-E6 cell line was derived). Note that overall the ACE2 proteins are highly similar to each other. The alignment and image were produced by Jalview.

**Fig. S7.**
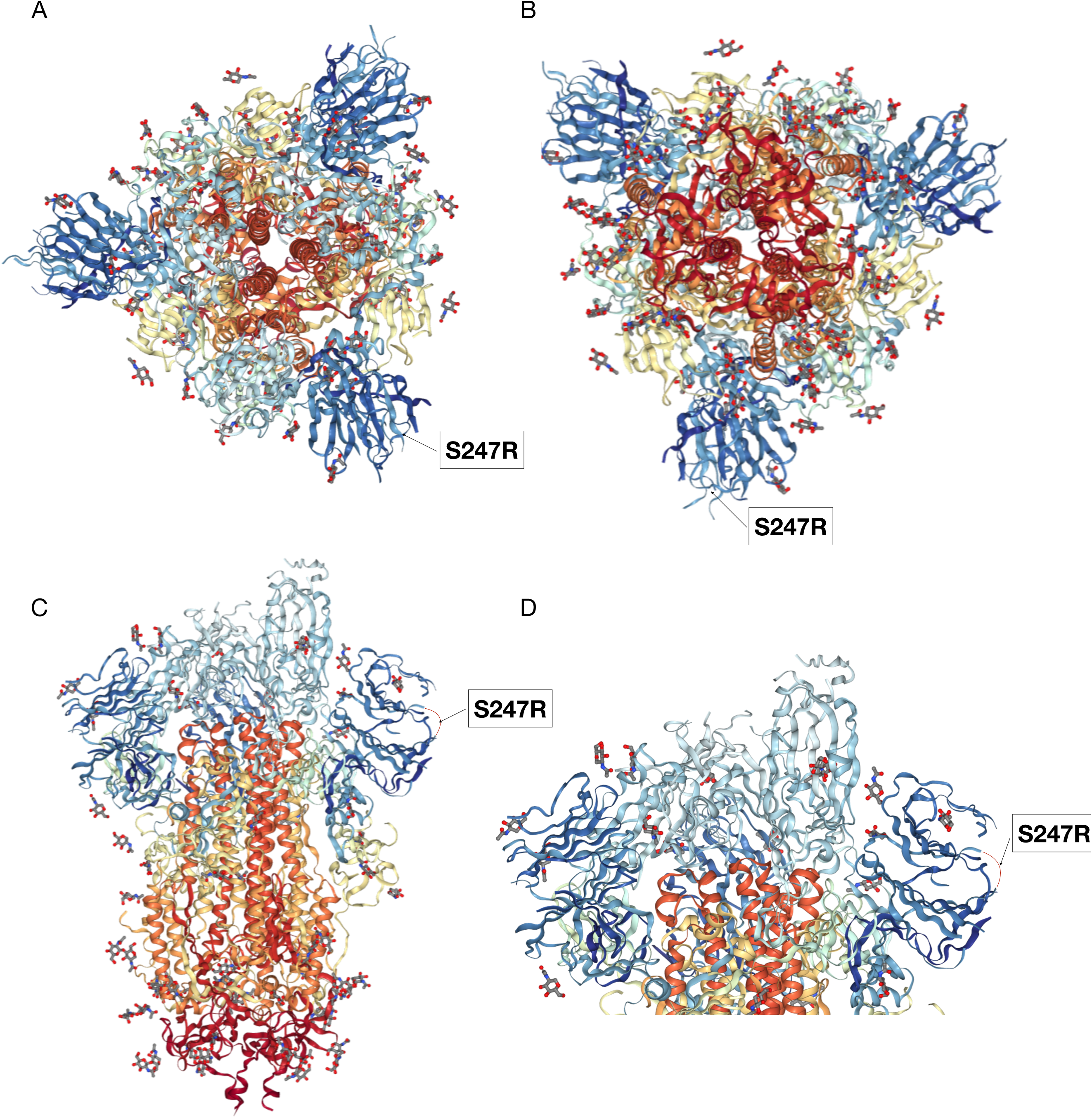
The 3D structure of the S-protein with the S247R overlay. The top (A), bottom (B), side (C), and close-up view (D) were provided. Note that the actual position of S247 was not determined in the original structure, hence a small red arc was in place to represent to the potential flexible loop conformation for (C) and (D). Also note that the protein complex is trimeric, but only one of the three mutations was labeled. The 3D structure of the S protein was visualized and downloaded from https://www.rcsb.org/3d-view/6VSB/1.

**Fig. S8.**
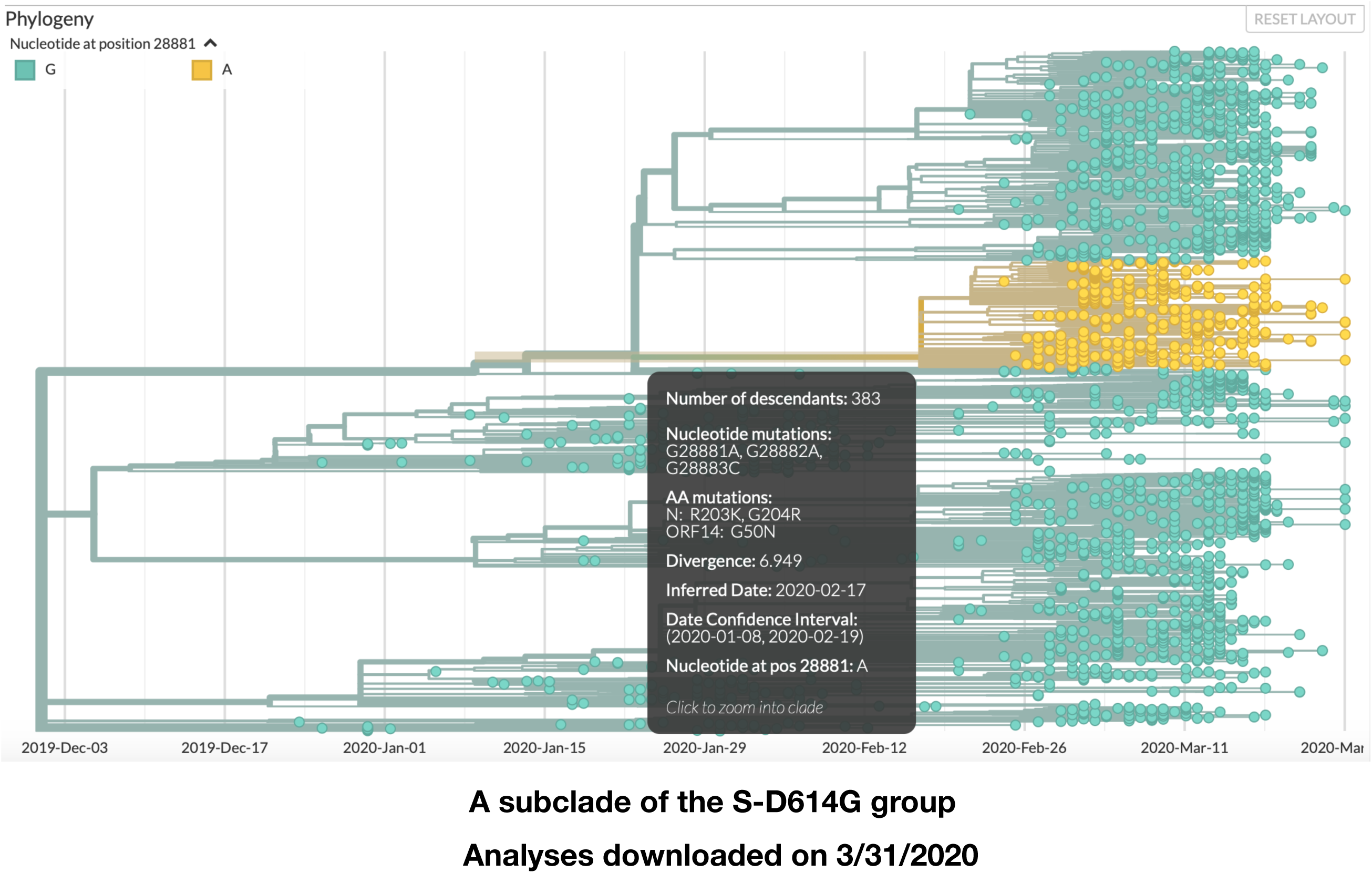
The trinucleotide mutation (G28881A, G28882A, and G28883C) was identified in the GISAID dataset and is the founding mutation for a large cluster of viral isolates within the S-D614G group (European clade).

## Author Contributions

M.Z., C.J., N.W., and L.L conceived and supervised the study. H.Y. and X.L. performed all the experiments, with help from K.X., Y.C., L.C., F.L., Z.W., H.W., and C. Jin. C.J., M.Z., and Q.C. performed all the data analyses. C.J., Q.C., H.Y., and M.Z. drafted and revised the manuscript with input from all authors.

## Acknowledgments

We gratefully acknowledge Drs. X. Zhu, L. Xiang, J. Jensen, and M. Lynch for their helpful discussions. We thank Dr. V. Billing for her help on improving the manuscript.

